# NK Cell Activation as an Immune Mechanism in Depression: Insights from Single-Cell Multi-Omics

**DOI:** 10.64898/2026.10.23.26366302

**Authors:** Matthew Boyton, Atsushi Takata, Fergus Hamilton, Robert Dantzer, Edward T Bullmore, Tom R Gaunt, Golam M Khandaker

## Abstract

Major depressive disorder (MDD) is associated with systemic immune dysregulation, but the specific immune cell populations and activation states contributing to disorder susceptibility remain poorly understood. Identifying these dynamic immune mechanisms may help uncover new opportunities for immune-targeted interventions in depression. We combined single-cell transcriptomic data from peripheral blood mononuclear cells (PBMCs) under resting and stimulated conditions with population genetic analyses to identify immune-cell populations and activation states linked to MDD susceptibility. Psoriasis was included as a comparator phenotype where ground truth regarding immune involvement is established. We identified immune-cell programmes enriched for disease-associated genetic risk and used Mendelian randomisation to prioritise genes potentially contributing to disease susceptibility. We find that psoriasis heritability predominantly localised to activated lymphocyte populations following CD3/CD28 stimulation, consistent with its established adaptive immune pathology. In contrast, MDD heritability was enriched within natural killer (NK) cell populations among resting and lipopolysaccharide (LPS)-stimulated PBMCs. Genetic analyses further prioritised NK-cell-associated genes, including *CCL5*, *GZMA* and *SYNE2*, with evidence of genetically predicted effects on MDD risk. By connecting genetic evidence for immune involvement in depression with specific cellular and functional contexts, our findings implicate NK-cell transcriptional programmes in MDD susceptibility, highlighting NK-cell immune signalling as a potential target for immunomodulatory treatment strategies.

## Introduction

Major depressive disorder (MDD) is a leading cause of health related disability^1^. Converging epidemiological, clinical and molecular evidence has implicated inflammatory processes in the pathophysiology of MDD^2, 3^. Patients with MDD consistently demonstrate alterations in circulating inflammatory markers, including C-reactive protein (CRP), interleukin-6 (IL-6), tumour necrosis factor-α (TNF-α), and interleukin-1β (IL-1β)^4–7^, while elevated inflammatory burden has been associated with symptom severity, treatment resistance, and poorer clinical outcomes^8, 9^. More recently, studies have demonstrated that these effects represent opportunities for therapeutic intervention, including evidence that IL-6 blockade improves multidomain symptoms in difficult-to-treat depression among individuals with elevated inflammatory biomarkers^10^. In parallel, depression is more common in people with chronic inflammatory disease characterised by altered immune regulation, such as psoriasis and rheumatoid arthritis^11–14^. Genetic studies suggest that these somatic and psychiatric conditions share components of their underlying biological architecture^15^. Therefore, it is plausible that immune regulation represents a biologically meaningful and therapeutically relevant component of MDD pathology.

A crucial question remains as to the specific cellular mechanism through which immune dysregulation contributes to MDD risk. Immune cells exist along a dynamic continuum of functional states that are shaped by environmental stimulation, differentiation and inter- and intra-cellular signalling^16^. However, much of the current literature has relied on circulating cytokine measurements, bulk transcriptomic profiling, or conventional genome-wide association studies (GWAS), each of which provides a partial view of dynamic immune biology and treats immunological processes as static biological features. Furthermore, bulk molecular approaches average signals across heterogeneous immune cell populations and therefore cannot determine which specific immune cell types contribute to disease-associated transcriptional changes. These limitations highlight the need for analytical approaches capable of resolving dynamic cell-type specific effects with genetic causal inference methods^17^. Identifying the immune cell populations and activation states through which these effects operate could ultimately inform more precise therapeutic targeting for greater efficacy.

Understanding how genetic susceptibility for disease maps onto dynamic immune states may provide important mechanistic insight into the biological processes underpinning conditions such as MDD. Activation state represents a fundamental determinant of immune cell function^18^. Following exposure to inflammatory stimuli, immune cells undergo transcriptional reprogramming and coordinated changes in signalling pathways, effector functions and cellular interactions^19, 20^. These responses differ substantially between innate and adaptive immune activation, reflecting distinct biological roles in host defence and immune regulation^21^. As a result, transcriptional programmes identified under resting conditions may differ markedly from those induced following immune stimulation, with activation-specific gene networks potentially capturing disease-relevant biology that is unobservable under baseline conditions^22^.

Advances in single cell transcriptomics have provided a technology by which dynamic immune activation processes can be queried at scale^23^. Experimental stimulation of peripheral blood mononuclear cells (PBMCs) enables the characterisation of coordinated transcriptional responses across diverse immune lineages^19^. When integrated with statistical genetic approaches, such as heritability enrichment and Mendelian randomisation (MR)^24^, these datasets provide a powerful framework for linking disease-associated genetic variation to specific immune activation states and causal molecular drivers of disease.

In this study, we combined single-cell transcriptomic data from stimulated PBMCs with GWAS summary statistics to identify peripheral immune cell and activation-state-specific mechanisms contributing to MDD susceptibility (**Figure 1**). Psoriasis, a well-characterised immune-mediated disease with established adaptive immune biology, was included as a comparator phenotype to evaluate the ability of this framework to reveal known disease mechanisms. Finally, we prioritised molecular drivers of disease within dynamic cellular activation signatures by identifying cell-type and stimulation-state-specific transcriptional programmes and applying MR to candidate molecular exposures.

**Figure 1.**
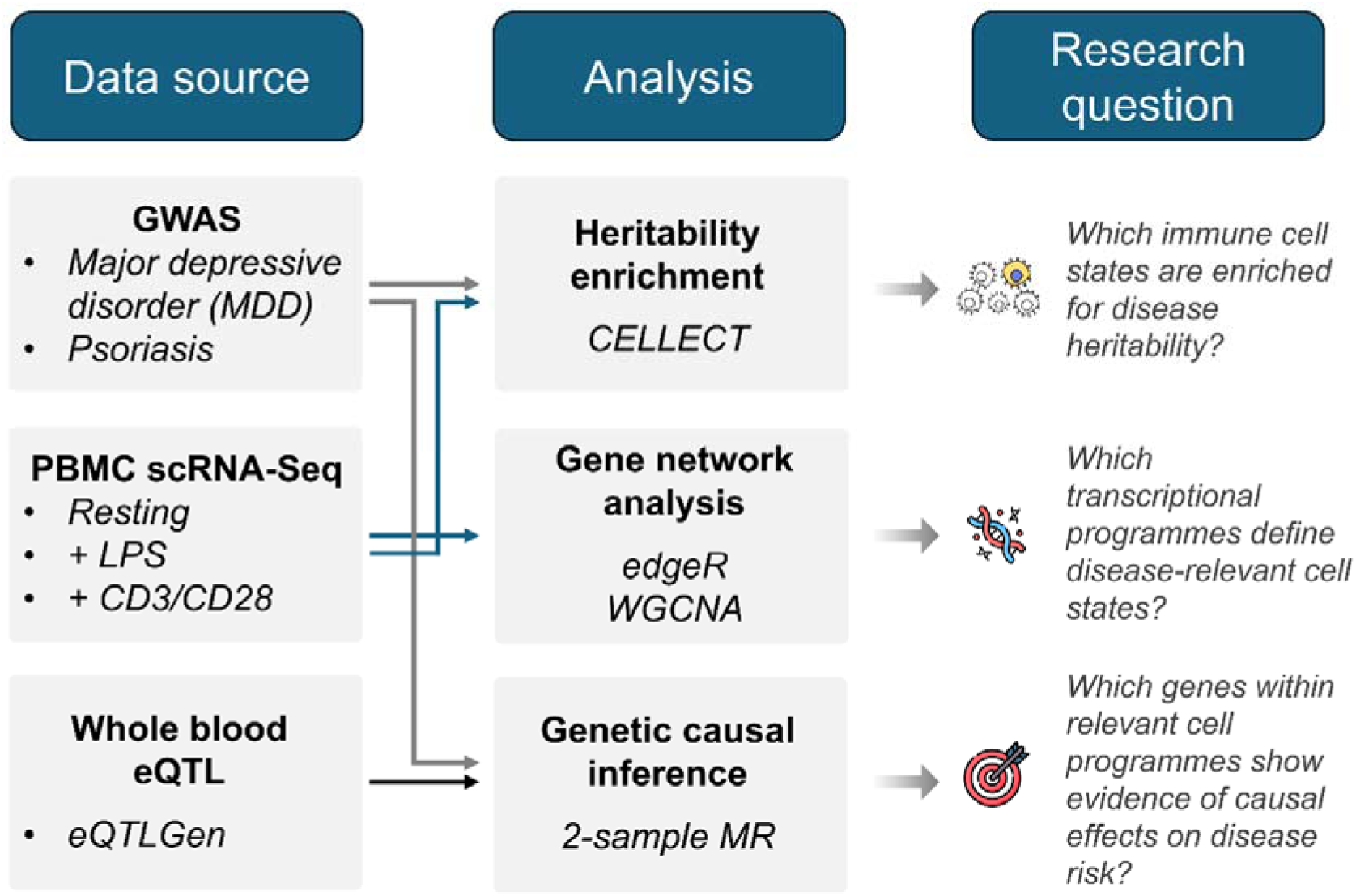
Integrative-multi-omic framework for identifying dynamic molecular immune drivers of disease risk. The framework integrates population genetic data (GWAS), stimulated single-cell transcriptomic data (scRNA-seq), and expression quantitative trait loci (eQTLs), first to identify disease-relevant immune cell states and transcriptional programmes, and subsequently to prioritise molecular exposures through 2-sample Mendelian randomisation. This approach provides cell-type and activation-state context for genetically supported molecular drivers of disease risk. Abbreviations: CD3/CD28, cluster of differentiation 3/28; CELLECT, CELL-type Expression-specific integration for Complex Traits; eQTL, expression quantitative trait locus; GWAS, genome-wide association study; LPS, lipopolysaccharide; MDD, major depressive disorder; MR, Mendelian randomisation; PBMC, peripheral blood mononuclear cell; scRNA-seq, single-cell RNA sequencing; WGCNA, weighted gene co-expression network analysis

## Methods

### Computational workflow and code availability

**Figure 1** shows an overview of the analytic pipeline. All analyses were conducted using R version 4.3.3 and Python version 3.11.9 on the University of Bristol’s high-performance computing cluster.

Specific code, formatted data resources, and implementation details are available at the publication’s GitHub repository: https://github.com/mboyton/pbmc-transcriptomics-heritability-mdd.

### PBMC stimulation atlas

Single-cell RNA sequencing and CITE-seq data, which enables simultaneous profiling of gene expression and cell-surface proteins, were obtained from the PBMC stimulation atlas generated by Lawlor et al^19^. This dataset was selected because it profiles multiple immune cell populations across resting, innate and adaptive stimulation conditions, enabling cell-type-specific transcriptional responses to distinct immune activation states to be compared. PBMCs were isolated from ten healthy young adult donors (five male, five female) and cultured under three experimental conditions: resting baseline state, innate immune activation via lipopolysaccharide (LPS) stimulation, and adaptive immune activation via anti-CD3/CD28 stimulation. The original dataset comprised 16,382 single cells spanning major peripheral immune populations, including B cells, CD4+ and CD8+ T cells, monocytes and NK cells. Cells were profiled using multiplexed CITE-seq, simultaneously quantifying transcriptomic and cell-surface protein expression.

Processed expression matrices and accompanying cell annotations provided by the original study were used for all downstream analyses. Cells lacking a cell-type annotation were excluded prior to analysis. To generate activation-state-specific immune populations, published cell-type annotations were combined with experimental stimulation-condition labels to create unique cell-state annotations representing seven immune cell lineages across three experimental conditions (21 cell states in total). These activation-state-specific populations were used for all downstream cell-state heritability enrichment analyses and transcriptional programme prioritisation.

### GWAS datasets

Summary statistics for MDD were obtained from the most recent Psychiatric Genomics Consortium GWAS^25^. The study included 688,808 MDD cases and 4,364,225 controls recruited across multiple international cohorts and ancestries.

Summary statistics for psoriasis were obtained from the GWAS meta-analysis reported by Dand et al^26^, comprising 36,466 psoriasis cases and 458,078 controls of predominantly European ancestry across 18 contributing GWAS cohorts.

### GWAS harmonisation and formatting

Raw summary statistics were reformatted into a standardised GWAS structure prior to downstream analyses. Variant identifiers, effect alleles, non-effect alleles, effect estimates, standard errors, association p-values, and sample size information were retained where available. Z scores were calculated as β/SE and summary statistics were formatted into a CELLECT/LDSC-compatible structure containing SNP, effect allele (A1), non-effect allele (A2), effect estimate (β), standard error (SE), p-value (P), sample size (N), and Z-score fields.

For the psoriasis GWAS, variants were additionally annotated with dbSNP reference identifiers using a GRCh37 reference panel^27^. Where rsIDs were unavailable, chromosome-position-allele identifiers were used to ensure complete variant annotation.

The resulting formatted summary statistics were used for both cell-state heritability enrichment analyses and MR.

### Cell-state heritability enrichment

#### Generation of cell-state specificity matrices (CELLEX)

Cell lineage and activation-state-specific expression specificity (ESμ) matrices were generated using single-cell transcriptomic profiles from the Lawlor et al. stimulation atlas via CELLEX^28^. ESμ scores were calculated for each gene across activation-state-specific immune populations, quantifying the relative specificity of gene expression within each cell state.

#### Heritability enrichment analyses (CELLECT)

Expression specificity matrices generated via CELLEX and formatted GWAS summary statistics underwent heritability enrichment analyses using CELLECT^28^. CELLECT integrates cell-state-specific expression specificity scores with GWAS summary statistics using stratified linkage disequilibrium score regression (LDSC) to estimate the contribution of individual cell states to trait heritability. Enrichment analyses were performed independently across all resting and stimulated PBMC populations (21 activation-state-specific immune populations in total). Cell states demonstrating positive regression coefficients with nominal evidence of enrichment (*P* < 0.05) were prioritised for downstream transcriptional programme analyses.

#### Resting-state transcriptional signatures

Resting-state transcriptional signatures were identified using donor-level pseudobulk differential expression analyses implemented in the *edgeR* package^29^. For each immune lineage, cells from the resting condition were grouped into the focal cell type and all remaining resting PBMC populations. Raw gene counts were summed within each donor-by-group combination to generate one pseudobulk expression profile per donor and comparison group. Genes with insufficient expression were removed using *filterByExpr*, after which library sizes were normalised using the trimmed mean of M-values method. Quasi-likelihood generalised linear models were fitted with cell group and donor included as model terms, thereby accounting for inter-individual variation while testing for lineage-specific expression differences. Differential expression was assessed by contrasting each focal resting immune population against all other resting PBMC populations. Genes demonstrating positive differential expression (false discovery rate (FDR) *P* < 0.05 and log_2_ fold-change > 1) were retained as resting-state marker genes for downstream MR.

#### Activation-state transcriptional programmes

Activation-associated transcriptional programmes were identified separately for each immune lineage and stimulation contrast using weighted gene co-expression network analysis (WGCNA), implemented via the *WGCNA* R package^30^. Lineage-specific pseudobulk expression matrices were analysed with samples representing individual donors under resting and stimulated conditions. Genes and samples failing the *goodSamplesGenes* quality checks were removed, after which the top 50% most variable genes were retained for network construction.

Signed co-expression networks and signed topological overlap matrices were constructed. Candidate soft-thresholding powers of 1–10, 12, 14, 16, 18 and 20 were evaluated, and the lowest power achieving a signed scale-free topology fit of R² ≥ 0.80 was selected. Where no candidate reached this threshold, the power with the highest signed R² was used. Modules were detected using hierarchical clustering and dynamic tree cutting, with a minimum module size of 50 genes, and modules with highly similar eigengenes were merged using a cut height of 0.25.

Module eigengenes were correlated with activation status, encoded as resting = 0 and stimulated = 1. Module–trait association *P*-values were corrected using FDR, and modules with FDR < 0.05 were retained as activation-associated transcriptional programmes. For each gene, module membership was quantified using eigengene connectivity (kME), and mean expression differences between stimulated and resting conditions were calculated. These statistics were used to prioritise highly connected genes within disease-relevant activation modules for downstream MR.

#### Mendelian randomisation integration with disease-associated transcriptional programmes

To estimate the causal effects of gene expression programs on risk of MDD or psoriasis, we applied 2-sample MR using the *TwoSampleMR* R package (version 0.6.7)^31^. MR inference assumes that genetic instruments are associated with the exposure, are independent of confounders, and influence the outcome only through the exposure^24^. Gene expression quantitative trait loci (eQTLs) were obtained from eQTLGen^32^. Transcriptome-wide MR analyses were first performed across all available eQTLGen gene-expression exposures using MDD and psoriasis as outcome phenotypes. The resulting estimates were subsequently integrated with transcriptional programmes identified within disease-relevant immune cell states, avoiding repeated estimation of identical gene–disease MR estimates across overlapping programmes.

Exposure and outcome datasets were harmonised to ensure that effect alleles were aligned to the exposure effect allele, such that causal estimates represent the change in disease risk per standard deviation increase in gene expression. Variants unavailable or excluded during harmonisation were omitted from MR analyses. Genetic instruments were defined as genome-wide significant (p < 5×10^-8^) cis-QTL variants, located within 1Mb of the gene centre and clumped for linkage disequilibrium at r² < 0.001 using the default PLINK-based procedure implemented in *TwoSampleMR* with reference to the 1000 Genomes Project Phase 3 European ancestry reference panel^33^.

For resting cell states, MR-tested genes were intersected with lineage-enriched markers identified using *edgeR*. For stimulated cell states, MR-tested genes were intersected with genes assigned to WGCNA modules associated with activation.

Genes demonstrating nominal evidence of causal effect in inverse-variance weighted (IVW) MR (*P* < 0.05) were retained for reporting and visualisation.

Reporting followed the STROBE-MR guidelines for Mendelian randomisation studies^34^. The completed checklist is provided in **Supplementary File 1**.

## Results

### Heritability enrichment highlights disease risk within distinct immune cell activation states

To identify immune activation states enriched for disease heritability, we applied CELLECT^28^ using resting and stimulated PBMC transcriptomic profiles with GWAS summary statistics for psoriasis and MDD.

Psoriasis heritability predominantly localised to adaptive lymphocyte activation states, with additional enrichment in activated NK-cell programmes. The strongest enrichment signals were observed in CD3/CD28-stimulated CD8+ memory T cells (*P* = 0.0010) and CD4+ memory T cells (*P* = 0.0013), both of which remained significant after correction across the 21 tested cell states. Additional nominal enrichment was observed in CD3/CD28-stimulated CD4+ naive T cells (*P* = 0.010), CD8+ naive T cells (*P* = 0.043), and NK cells from stimulated PBMC cultures (*P* = 0.016) (**Figure 2A**; **Supplementary Table 1**). Activated NK-cell programmes also showed nominal enrichment following LPS stimulation of PBMC cultures (*P* = 0.039).

**Figure 2.**
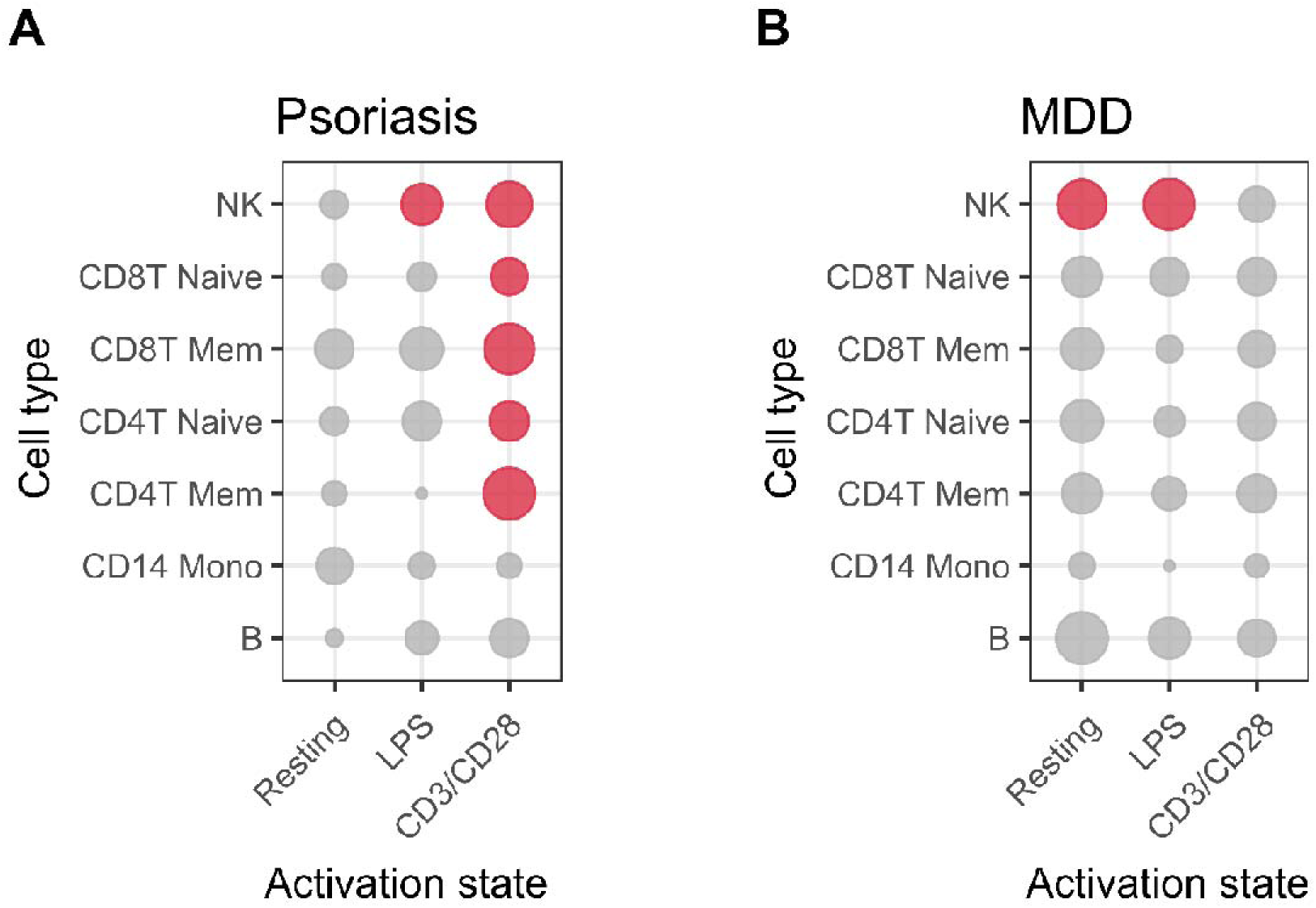
Psoriasis and major depressive disorder (MDD) exhibit distinct immune cell-state heritability enrichment profiles. CELLECT was applied to estimate GWAS heritability among resting and stimulated PBMC transcriptional programmes. Point size indicates the magnitude of the heritability enrichment coefficient, red points indicate evidence of nominal heritability enrichment (*P* < 0.05).

In contrast, we observed a comparatively NK-cell-focused enrichment profile in MDD, spanning both resting and innate stimulation states. Enrichment was observed in resting NK cells (*P* = 0.042) and in NK cells within LPS-stimulated PBMC cultures (*P* = 0.040), whereas no comparable evidence was observed among CD3/CD28-stimulated populations (**Figure 2B**; **Supplementary Table 1**). These signals were treated as nominal evidence used to guide subsequent transcriptional programme analyses.

### Activation-associated transcriptional programmes identify disease-relevant immune modules

Following identification of disease-associated immune activation states, we defined resting and activation-associated transcriptional programmes within each prioritised lineage. Resting cell-state signatures were identified using donor-blocked quasi-likelihood GLM differential expression analyses comparing each resting lineage against all other resting PBMC populations (**Supplementary Table 2**). Activation-associated transcriptional programmes were identified independently within each lineage using WGCNA, with modules associated with stimulation identified from module eigengene–trait correlations (**Supplementary Table 3**). Highly connected genes (kME ≥ 0.75) from activation-associated modules, together with lineage-enriched resting markers, were subsequently prioritised for Mendelian randomisation analyses (**Supplementary Table 4**).

### Psoriasis-associated activation modules contain risk altering gene programmes

To identify candidate causal genes within psoriasis-associated transcriptional programmes, we performed IVW MR using whole-blood cis-eQTL instruments from eQTLGen, restricting analyses to genes prioritised from activation-associated WGCNA modules within psoriasis-enriched immune cell states.

Multiple genes demonstrated evidence of altered psoriasis risk across CD4+, CD8+, and NK-cell activation programmes (**Figure 3**). Among the strongest effects, increased genetically predicted *UBE2L3* expression demonstrated evidence of increased psoriasis risk (OR = 1.15, 95% CI 1.12–1.19, *P* = 1.4 × 10^-18^), with the gene prioritised across CD3/CD28-responsive programmes from CD4+ memory, CD4+ naive, CD8+ memory, CD8+ naive, and NK cells. Increased genetically predicted *NFKB1* expression also demonstrated evidence of increased psoriasis risk (OR = 1.24, 95% CI 1.12–1.37, *P* = 3.9 × 10^-5^) and was prioritised across four CD4+ and CD8+ T-cell activation programmes. Conversely, increased genetically predicted expression of *HSPA4* (OR = 0.85, 95% CI 0.81–0.90, *P* = 6.2 × 10^-8^) and *BANF1* (OR = 0.87, 95% CI 0.83–0.92, *P* = 2.4 × 10^-6^) demonstrated evidence of reduced psoriasis risk, with both genes prioritised across multiple activation-associated programmes. Full MR results for genes within the prioritised psoriasis activation-associated transcriptional programmes are provided in **Supplementary Table 5**.

**Figure 3.**
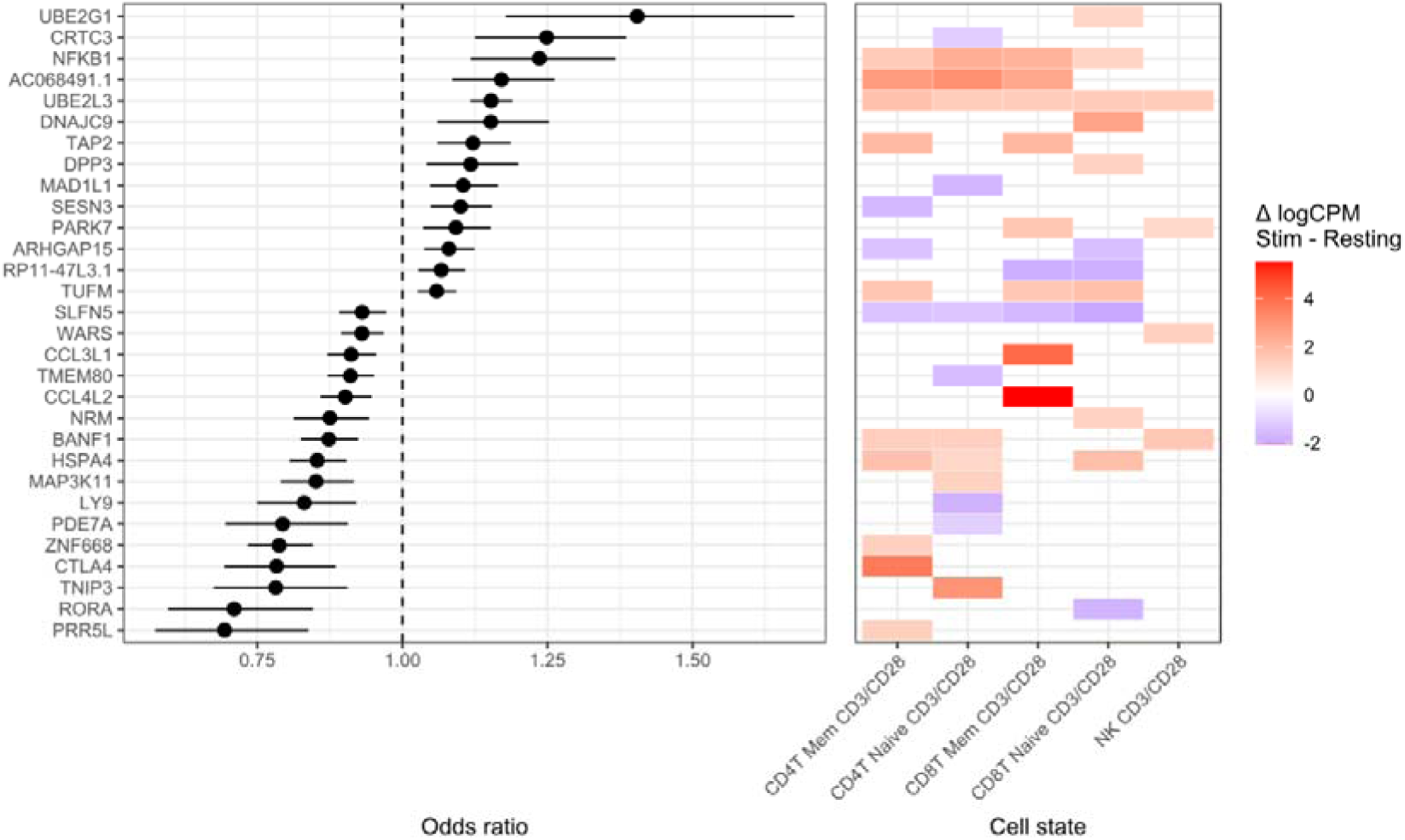
Mendelian randomisation prioritises genes within psoriasis-associated immune activation programmes. The left panel shows inverse-variance weighted Mendelian randomisation estimates (odds ratios and 95% confidence intervals) for the top psoriasis-associated genes prioritised from activation-associated transcriptional programmes. The right panel indicates the immune activation state(s) in which each gene was identified, with colour representing the direction and magnitude of differential expression following stimulation (ΔlogCPM; stimulated minus resting), where positive values indicate higher expression following stimulation and negative values indicate lower expression following stimulation relative to the resting state. Genes were prioritised from significant WGCNA modules (module FDR < 0.05; kME ≥ 0.75).

Several findings reflected established immune regulatory biology. Increased expression of *CTLA4* showed evidence of a protective effect in psoriasis (OR = 0.78, 95% CI 0.69–0.89, *P* = 9.0 × 10^-5^), despite being upregulated within activated lymphocyte programmes. This is consistent with its role as an immune checkpoint inhibitor; undergoing rapid upregulation following exposure to proinflammatory stimuli but propagating an anti-inflammatory effect in the longer term^35^. *RORA* similarly demonstrated evidence of reduced psoriasis risk (OR = 0.71, 95% CI 0.60–0.85, *P* = 1.3 × 10^-4^) and was prioritised within a CD8+ naive T-cell activation-associated programme. Additional prioritised genes included *CCL4L2*, *CCL3L1*, *TAP2*, *PDE7A*, *TNIP3*, and *PTPN2*, further highlighting dynamic pathways involved in lymphocyte activation and inflammatory signalling^36–38^.

Overall, these findings demonstrate that activation-state-informed transcriptional programmes reveal multiple established psoriasis susceptibility genes and immune regulatory pathways.

### Possible causal effects of NK-cell activation genes in MDD

Having identified established associations between immune cell transcriptional programs and disease risk for psoriasis, we extended our analytical framework to MDD. MR analyses showed potential causal effects of multiple genes within resting and activation associated NK-cell transcriptional programmes on MDD risk (**Figure 4**).

**Figure 4.**
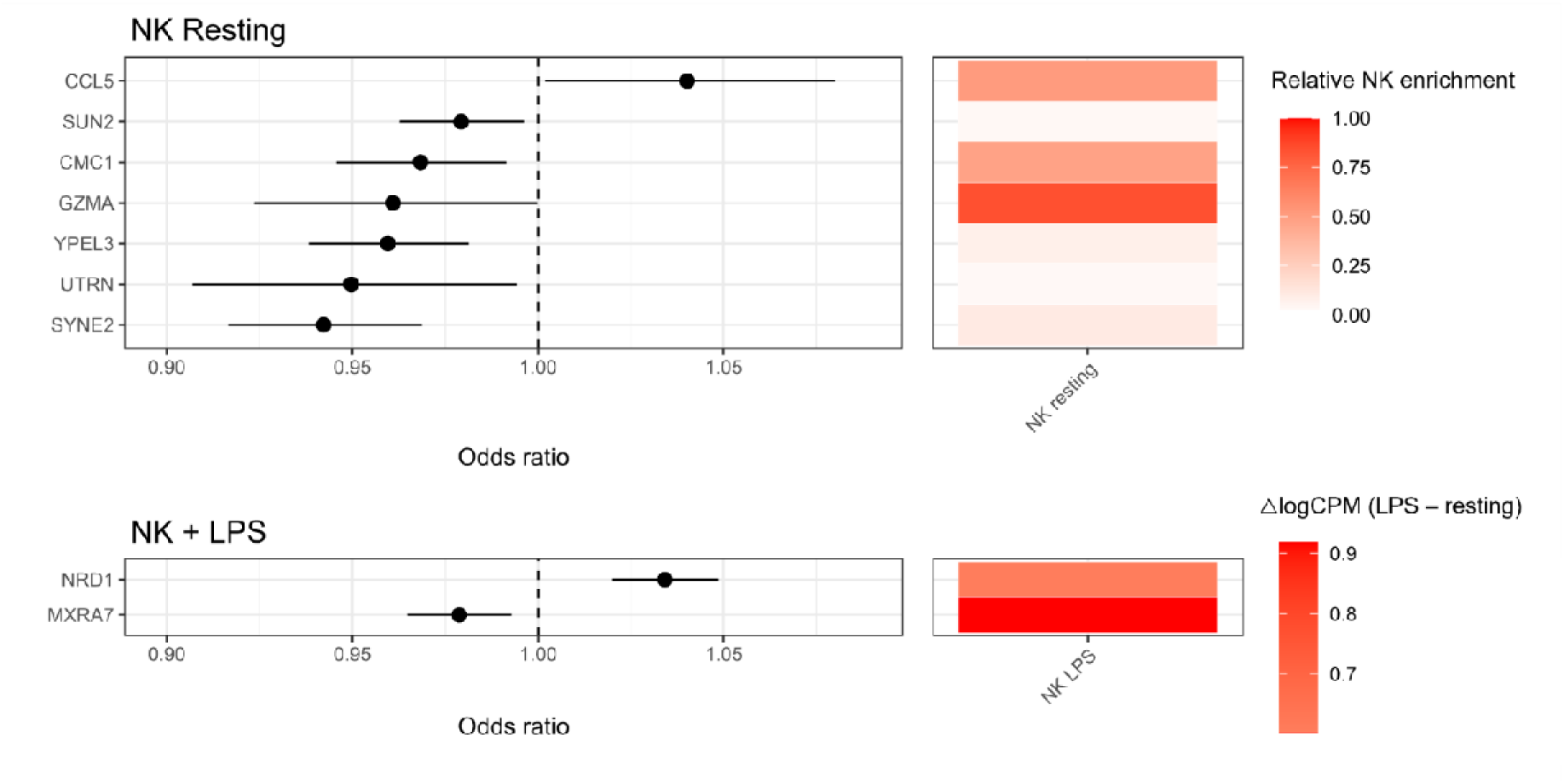
Mendelian randomisation prioritises candidate genes within MDD-associated NK-cell transcriptional programmes. The left panels show inverse-variance weighted Mendelian randomisation estimates (odds ratios and 95% confidence intervals) for the top MDD-associated genes prioritised from resting (top) and LPS-activation-associated (bottom) transcriptional programmes. The right panels indicate the immune activation state in which each gene was identified. Colours represent relative NK-cell enrichment for resting NK-cell genes (top), derived from differential expression effect sizes comparing resting NK cells with other resting PBMC populations, with higher values indicating greater relative enrichment. For LPS-responsive genes (bottom), colours represent the direction and magnitude of expression change following stimulation (ΔlogCPM; LPS-stimulated minus resting), with positive values indicating higher expression following LPS stimulation.

Within resting NK-cell programmes, increased genetically predicted expression of *CCL5* (OR = 1.04, 95% CI 1.00–1.08, *P* = 0.040) showed evidence for a risk-increasing effect on MDD. Conversely, several genes demonstrated evidence of protective effects, including *SYNE2*, *YPEL3*, *CMC1*, *SUN2*, *UTRN*, and *GZMA*. Of these, *SYNE2* (OR = 0.94, 95% CI 0.92–0.97, *P* = 2.6 × 10^-5^, FDR = 0.0013) and *YPEL3* (OR = 0.96, 95% CI 0.94–0.98, *P* = 3.1 × 10^-4^, FDR = 0.0076) showed evidence of genetic effects following FDR adjustment.

Within LPS-responsive NK-cell activation modules, increased genetically predicted expression of *NRD1* demonstrated evidence of increased MDD risk (OR = 1.03, 95% CI 1.02–1.05, *P* = 2.1 × 10^-6^, FDR = 3.3 × 10^-5^), while increased genetically predicted expression of *MXRA7* demonstrated evidence of reduced MDD risk (OR = 0.98, 95% CI 0.96–0.99, *P* = 0.0033, FDR = 0.027). Full MR results for genes within the LPS-responsive and resting NK-cell transcriptional programmes are provided in **Supplementary Tables 6 and 7**, respectively.

Sensitivity analyses using alternative MR estimators supported the direction of the primary IVW estimates for all four genes with ≥3 independent instruments (*SYNE2*, *CMC1*, *GZMA* and *MXRA7*). Weighted-median estimates additionally supported effects for *SYNE2*, *CMC1* and *MXRA7*, while estimates from the remaining methods were directionally concordant but less precise, consistent with the small number of available instruments (3 - 4 SNPs per gene; **Supplementary Figure 1**; **Supplementary Table 8**)

These findings prioritise a set of genes within both resting and activation-associated NK-cell transcriptional programmes, providing convergent evidence that NK-cell biology contributes to genetic susceptibility for MDD.

## Discussion

Our integrative multi-omic framework combined stimulation-aware single-cell PBMC transcriptomics with statistical genetics to identify immune activation states underlying disease susceptibility. By combining cell-state heritability enrichment, transcriptional programme prioritisation, and genetic causal inference using MR, we identified distinct immune architectures underlying MDD and psoriasis. Psoriasis genetic liability predominantly localised to activated adaptive immune populations, whereas MDD risk preferentially mapped to resting and LPS-responsive NK-cell programmes. Then using MR we prioritised potentially causal genes within these disease-associated transcriptional programmes, providing both cellular and activation-state context for interpreting disease-associated molecular mechanisms.

Our data for psoriasis provide important benchmarking within a strongly immune-driven condition. Psoriasis has a well-established adaptive immune pathogenesis involving T-cell activation and IL-23 biology^39, 40^. Consistent with this understanding, psoriasis heritability localised predominantly to activated CD4+, CD8+, and NK-cell populations following CD3/CD28 stimulation. Furthermore, MR prioritised multiple established psoriasis susceptibility genes, including *UBE2L3* and *NFKB1*, demonstrating that the framework successfully recovers known disease mechanisms^41, 42^. We additionally identified the inhibitory checkpoint protein *CTLA4*^41^ and the immune regulatory phosphatase *PTPN2*^43^. The prioritisation of *CTLA4* is particularly consistent with the activation-state context of our analytical framework, since *CTLA4* expression is strongly induced following T-cell activation and subsequently restrains CD28-mediated co-stimulation^44^. Its protective MR effect is consistent with its established role as an activation-induced negative regulator of T-cell inflammatory signalling^35^, and is further supported by the therapeutic targeting of the *CTLA4* co-stimulatory pathway by abatacept in psoriatic arthritis^45^. Alongside these established signals, we also prioritised emerging novel psoriasis genes including *BANF1*, previously implicated in inflammatory regulation in psoriatic keratinocytes^46^, and *HSPA4*, for which previous multi-omic analyses have provided preliminary evidence^47^. These findings provide genetic support for less well-characterised molecular pathways that may contribute to psoriasis susceptibility.

In contrast to the predominantly adaptive immune architecture observed for psoriasis, a key finding within this study is the enrichment of MDD genetic liability within NK-transcriptional programmes. Immune dysregulation is increasingly recognised as an important component of the biology of depression, with numerous studies reporting altered circulating cytokine concentrations, inflammatory biomarkers, and immune cell composition in affected individuals^2, 5, 6, 8–10, 48^. Much of this evidence has relied on bulk transcriptomic profiling or circulating biomarker measurements. By integrating stimulation-specific single-cell transcriptomics with genetic causal inference, our framework extends these observations by identifying both the immune cell lineage and functional activation state through which disease-associated genetic liability may be enriched. The observation that MDD heritability may localise to both resting and LPS-responsive NK-cell programmes suggests that NK-cell biology represents an important component of the peripheral immune architecture underlying depression susceptibility.

Our findings are consistent with extensive literature linking altered NK abundance and cytotoxic function with MDD^49, 50^, including a recent large-scale transcriptomic mega-analysis implicating NK-cell subsets in depression-associated gene expression differences^51^.

Importantly, NK cell function has been shown to recover following successful antidepressant treatment and symptom remission^52^, suggesting that alterations in NK-cell function reflect biologically relevant disease processes. Our findings complement these studies by demonstrating that genetic risk and resilience factors for MDD localise in transcriptional programmes within resting and LPS-responsive NK cells, providing genetic evidence linking observed immune abnormalities with inherited susceptibility.

LPS stimulation induces coordinated inflammatory responses within peripheral immune cells through Toll-like receptor (TLR) signalling, activating transcriptional programmes involved in inflammatory signalling and cellular stress responses^53^. Our findings suggest that inherited susceptibility to MDD preferentially maps to transcriptional programmes engaged during these innate immune activation states. Notably, the enrichment observed within LPS-responsive NK-cell programmes may reflect coordinated innate immune signalling rather than direct activation of NK cells by LPS itself. NK cells respond relatively weakly to LPS in isolation, with much of their activation occurring indirectly through cytokines released by monocytes and other innate immune cells following TLR stimulation^54^. Consequently, the transcriptional programmes identified here likely capture broad inflammatory communication networks acting upon NK cell biology, during states of innate immune activation.

Several genes prioritised through MR provide further insight into the biological processes linking NK-cell function with depression. Among LPS-responsive programmes, *NRD1* and *MXRA7* emerged as notable candidates. *NRD1*, a zinc-dependent metalloendopeptidase, has an established role in inflammatory signalling, including regulation of TNF-α pathways^55^. Previous studies have also linked its activity to various neurological conditions, including psychiatric phenotypes such as mood disorders^56^. The matrix-remodelling-associated protein *MXRA7* is involved in monocyte-to-macrophage differentiation^57^, and previous studies have linked its expression to postpartum depression^58^. Within resting NK-cell programmes, *CCL5* and *GZMA* were prioritised alongside *SYNE2*, *YPEL3*, *CMC1*, *SUN2*, and *UTRN*. *CCL5* and *GZMA* have particularly well-established roles in resting NK-cell biology, representing constitutive chemokine and cytotoxic effector components, respectively^59, 60^. CCL5–CCR5 signalling has additionally been implicated in MDD and related neuroimmune processes^61^, with elevated circulating CCL5 previously reported in affected individuals^62^. Interestingly, increased genetically predicted *GZMA* expression demonstrated evidence of reduced MDD risk. The remaining candidates implicate broader NK cell transcriptional processes, including nuclear-cytoskeletal architecture, mitochondrial metabolism and proliferation^63^. Taken together, these signals support the concept that genetically driven risk factors for MDD may converge within co-ordinated NK-cell transcriptional programmes.

This study also demonstrates the value of incorporating dynamic immune activation states into statistical genetic analyses to identify mechanism of complex disease more precisely. Conventional MR studies typically evaluate molecular exposures without considering the cellular context in which they are expressed, which can miss important condition-dependent genetic effects^64^. Immune transcriptional programmes change substantially following activation, and disease-associated genetic effects may only become apparent within particular functional states^22^. By integrating experimentally stimulated single-cell transcriptomics with heritability enrichment, co-expression network analyses, and MR, our framework enables disease-associated genetic variation to be traced from specific immune activation states through coordinated transcriptional programmes to candidate causal genes.

Several important limitations must be considered. First, the PBMC stimulation atlas was generated from a relatively small number of healthy donors and therefore may not capture the full diversity of immune activation responses present across populations, and may not fully reflect tissue-specific immune processes, such as brain or skin-specific mechanisms, underlying psoriasis and MDD risk^65^. Second, the stimulation experiments were restricted to LPS and CD3/CD28 activation among broad cell lineages, representing simplified models of innate and adaptive immune activation that do not encompass the full complexity of immune signalling among diverse lineages and subpopulations^66^. Third, both the stimulation atlas and the GWAS datasets were derived predominantly from individuals of European ancestry, limiting generalisability to other populations. Finally, IVW MR assumes that genetic instruments act independently through the exposure of interest, whereas genes within transcriptional programmes are likely highly correlated among shared biological pathways. Although individual MR effect estimates were modest, even small changes in disease risk per unit increase in genetically predicted gene expression may reflect biologically important immune signalling mechanisms.

In conclusion, our study demonstrates that stimulation-aware single-cell transcriptomic data can be integrated with genetic causal inference to identify activation-state specific mechanisms in psychiatric conditions. Integration of these complementary data sources revealed that genetic liability for MDD preferentially localises to NK cell transcriptional programmes, while simultaneously recovering known and novel biology in psoriasis. These findings provide further evidence that peripheral immune dysfunction contributes to MDD susceptibility and demonstrate the utility of dynamic immune-state context in improving the biological interpretation of genetic causal inference studies.

## Data Availability

All datasets analysed in this study were obtained from publicly available sources, including single-cell transcriptomic data, GWAS summary statistics and eQTL data. Results generated from these analyses are presented in the manuscript and accompanying supplementary materials.

https://www.frontiersin.org/journals/immunology/articles/10.3389/fimmu.2021.636720/full

https://www.eqtlgen.org/

https://www.ebi.ac.uk/gwas/studies/GCST90472771

https://pgc.unc.edu/for-researchers/download-results/

https://github.com/mboyton/pbmc-transcriptomics-heritability-mdd

## Acknowledgements

Matthew Boyton undertook part of this work while a visiting researcher in the Laboratory for Molecular Pathology of Psychiatric Disorders at the RIKEN Center for Brain Science, Japan. He gratefully acknowledges Atsushi Takata and his team for their support, collaboration and hospitality during his visit.

## Supplementary Figures

**Supplementary Figure 1.**
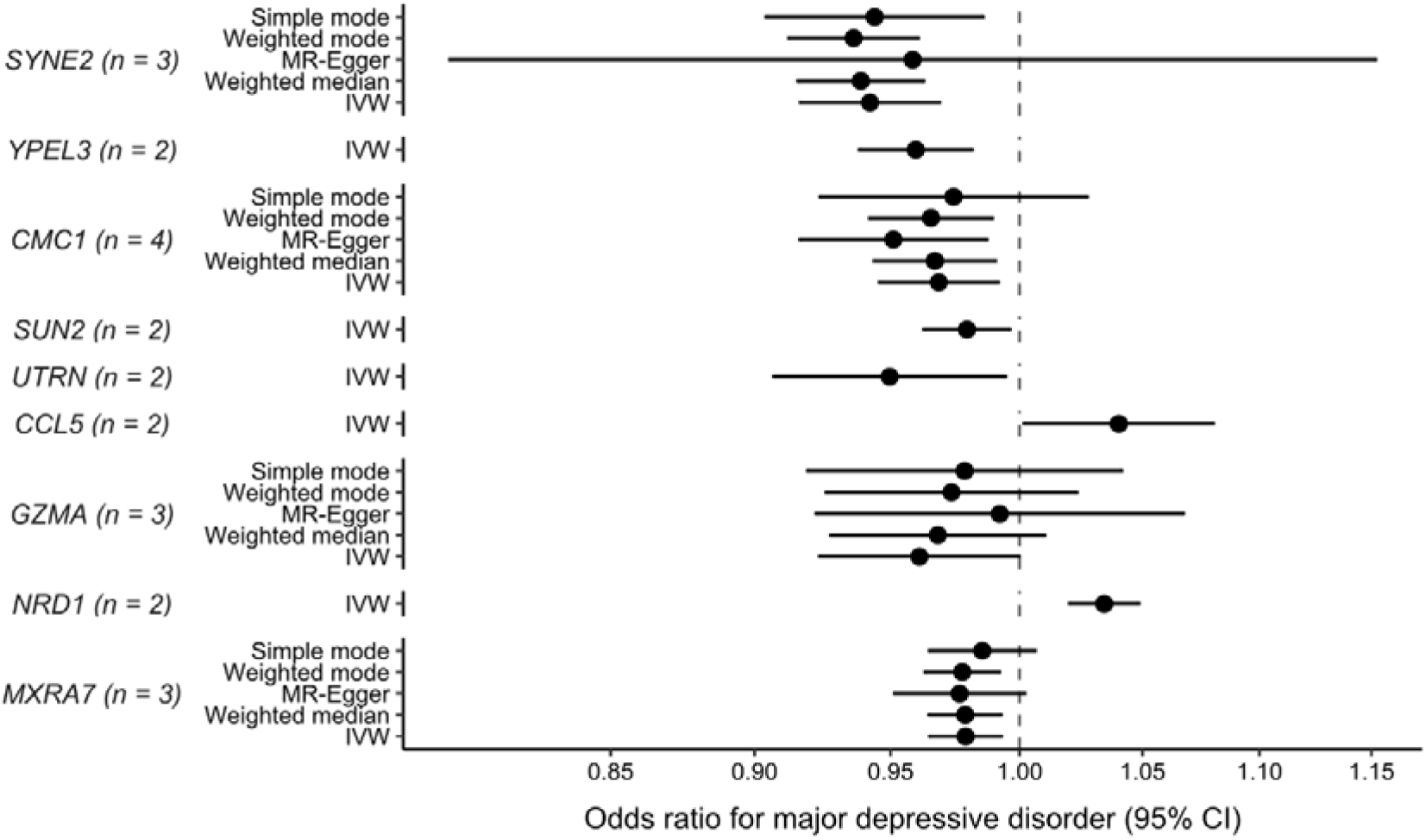
Sensitivity analyses of Mendelian randomisation estimates for NK-cell genes prioritised in major depressive disorder. Forest plots showing odds ratios (ORs) and 95% confidence intervals for major depressive disorder per genetically predicted increase in gene expression, estimated using inverse-variance weighted (IVW), MR-Egger, weighted median, simple mode and weighted mode methods. Genes are grouped by resting and LPS-responsive NK-cell transcriptional programmes. Alternative estimators are presented where at least three independent genetic instruments were available; genes with two instruments are represented by IVW estimates only. The number of instruments (n) is indicated alongside each gene. The dashed vertical line indicates the null effect (OR = 1)

